# Detection of pneumococcus during hospitalization for SARS-CoV-2

**DOI:** 10.1101/2022.07.13.22277607

**Authors:** Anne E. Watkins, Laura R. Glick, Isabel M. Ott, Samuel B. Craft, Devyn Yolda-Carr, Christina A. Harden, Maura Nakahata, Shelli F. Farhadian, Lindsay R. Grant, Ronika Alexander-Parrish, Adriano Arguedas, Bradford D. Gessner, Daniel M. Weinberger, Anne L. Wyllie

**Author notes:** co-senior authors (Daniel M. Weinberger, Anne L. Wyllie).

## Abstract

**Background:** Infections with respiratory viruses (e.g., influenza, RSV) can increase the risk of severe pneumococcal infections. Likewise, pneumococcal co-infection is associated with poorer outcomes in viral respiratory infection. However, there are limited data describing the frequency of pneumococcus and SARS-CoV-2 co-infection and the role of co-infection in influencing COVID-19 severity.

**Methods:** The study included patients admitted to Yale-New Haven Hospital who were symptomatic for respiratory infection and tested positive for SARS-CoV-2 during March-August 2020. Patients were tested for pneumococcus through culture-enrichment of saliva followed by RT-qPCR (to identify carriage) and serotype-specific urine antigen detection (UAD) assays (to identify presumed lower respiratory tract pneumococcal disease).

**Results:** Among 148 subjects, the median age was 65 years; 54.7% were male; 50.7% had an ICU stay; 64.9% received antibiotics; 14.9% died while admitted. Pneumococcal carriage was detected in 3/96 (3.1%) individuals tested by saliva RT-qPCR. Additionally, pneumococcus was detected in 14/127 (11.0%) individuals tested by UAD, and more commonly in severe than moderate COVID-19 (OR: 2.20; 95% CI: [0.72, 7.48]); however, the numbers were small with a high degree of uncertainty. None of the UAD-positive individuals died.

**Conclusions:** Pneumococcal LRTI, as detected by positive UAD, occurred in patients hospitalized with COVID-19. Moreover, pneumococcal LRTI was more common in those with more serious COVID-19 outcomes. Future studies should assess how pneumococcus and SARS-CoV-2 interact to influence COVID-19 severity in hospitalized patients.

**One Sentence Summary:** Pneumococcal lower respiratory tract infection, as detected by positive UAD, occurred in patients hospitalized with COVID-19 at rates similar to those reported prepandemic.

## INTRODUCTION

Most infections with SARS-CoV-2 are mild-to-moderate in severity while some progress to severe manifestations of COVID-19 including pneumonia, acute respiratory distress syndrome (ARDS), and death.^1^ The factors that determine disease severity remain incompletely understood. For other viral pathogens, co-infections with bacteria influence the severity of infection. This is evident during seasonal epidemics of influenza and respiratory syncytial virus (RSV), as well as during historical pandemics.^2,3^ *Streptococcus pneumoniae* (pneumococcus) is one of the most common co-infecting pathogens with influenza and RSV.^4^ There has therefore been interest in the possible role of bacterial co-infections in influencing the severity of COVID-19. Clinical studies have generally reported a small percentage of COVID-19 cases with secondary bacterial pneumonia.^5–8^ In contrast, the rates of COVID-19 disease, hospitalization, and death were lower among those who received pneumococcal conjugate vaccines (PCVs) compared with those who did not.^9–11^ This is consistent with previous work from a randomized trial that demonstrated lower rates of hospitalization for a number of viruses among children receiving PCVs, including endemic coronaviruses.^12^

We sought to quantify the prevalence of pneumococcal infection among patients hospitalized with COVID-19 and to explore interactions between pneumococcus and SARS-CoV-2 in patients hospitalized with COVID-19, including whether pneumococcal carriage or infection was associated with disease severity. We hypothesized that pneumococcus would be detected frequently in this population and that individuals co-infected with pneumococcus and SARS-CoV-2 would have more severe COVID outcomes compared with individuals infected with SARS-CoV-2 alone. We used data and samples from a retrospective cohort of individuals hospitalized with COVID-19 to evaluate the prevalence of pneumococcus and to explore the association with severity. Understanding these relationships is important when designing public health response measures that could mitigate both pneumococcal disease and COVID-19.

## MATERIALS AND METHODS

### Study design

During March-August of 2020, paired saliva and urine samples were collected from hospitalized patients at Yale-New Haven Hospital who were admitted to the hospital with symptomatic SARS-CoV-2 infection and enrolled in the Yale IMPACT study.^13^ Signed informed consent was obtained from all study participants following Yale University HIC-approved protocol #2000027690. Demographic and case information was collected through systematic and retrospective review of patient electronic medical records, including vaccination history and a peak measure of disease severity as described elsewhere.^14^ SARS-CoV-2 positive individuals with non-COVID reasons for admission (i.e. patients admitted for injury, to give birth, etc) and those under 18 years of age were excluded from this study.

### Sample collection

Pneumococcus is commonly found in the upper respiratory tract of healthy children^15^ and adults^16^ and can be detected by bacterial culture or qPCR in samples from the upper respiratory tract. Some individuals go on to develop lower respiratory tract infections (LRTIs). Because samples are not typically obtained from the lower respiratory tract, evidence of infection is indirect. The detection of pneumococcal capsular polysaccharides in the urine has been demonstrated to be a reliable marker of LRTIs caused by pneumococcus and can distinguish between carriage and disease.

Saliva self-collection was attempted every three days using previously described methods.^17^ Samples were transported to the Yale School of Public Health at room temperature within 5 hours of sample collection and tested for SARS-CoV-2 within 12 hours of sample collection. Urine collection was also attempted every three days from enrolled participants and stored at -80°C until further processing.

### Detection of pneumococcal carriage using the molecular method

For all COVID-19 inpatients with at least one SARS-CoV-2 positive test, remaining saliva samples were tested for pneumococcal carriage. Thawed saliva samples were first culture-enriched with 100 μ plated on gentamicin (10%) supplemented blood agar plates.^18^ Cultures were incubated overnight after which all growth was harvested and stored at -80°C.^19^ Pneumococcal detection was performed by DNA extraction of 200 μl of each sample using the MagMAX Ultra Viral/Pathogen Nucleic Acid Isolation kit (ThermoFisher Scientific) on the KingFisher DNA extraction robot (ThermoFisher Scientific) following manufacturer’s protocol. Samples were classified as positive when both *piaB*^19,20^ and *lytA*^21^ targets reported a cycle threshold (Ct) value <40 Ct by RT-qPCR.

### Detection of pneumococcal LRTI using Urine antigen detection (UAD)

Urine samples were thawed at room temperature and aliquoted into PIPES buffer. Aliquots were re-frozen at -80°C in preparation for batch shipping on dry ice to the reference laboratory of Pfizer Vaccine Research (Pearl River, NY) for testing.^22^ Upon receipt, samples were stored at -80°C until batched urine antigen testing could be performed using the serotype-specific UAD (SSUAD) and/or BinaxNOW tests according to manufacturer’s protocol.

### Statistical analysis

Differences in demographic data between pneumococcal status groups were tested using the Kruskal-Wallis rank sum test (continuous variable) or Fisher’s exact test (categorical variable). Estimates were considered statistically significant at *p*<0.05. All statistical analyses were performed in RStudio v1.2.133535, using R v3.6.1.36.

## RESULTS

### Study characteristics

Of the COVID-19 inpatients enrolled into the study, 156 saliva samples from 96 inpatients and 219 urine samples from 127 inpatients were available for testing (summarized in Table 1). Overall, data from 148 inpatients were included in the analysis in this study. Patient characteristics are summarized in Table 2. The overall cohort of SARS-CoV-2 positive participants included in this study was 54.7% male, with average age 64.6 years; 69 (46.6%) individuals were categorized as having severe COVID-19 disease, based on supplemental oxygen level requirements and admission to ICU.^14^ Antibiotic use at any point during hospitalization was high amongst all groups and was reported for 96/148 (64.9%; range: 64.1-71.4%) total inpatients. Ten patients had no bacterial growth following culture-enrichment of saliva; 8/10 (80.0%) of these patients had received antibiotics.

**Table 1.** Distribution of patient samples tested either by UAD and/or RT-qPCR of culture-enriched saliva.

|  | UAD | Saliva |
| --- | --- | --- |
| Total Inpatients | 127 | 96 |
| Total Samples | 219 | 156 |
| Positive Inpatients | 14 (11%) | 3 (3.1%) |
| 75/127 patients tested by UAD were also tested by saliva (none tested positive by both) |  |  |

**Table 2.**
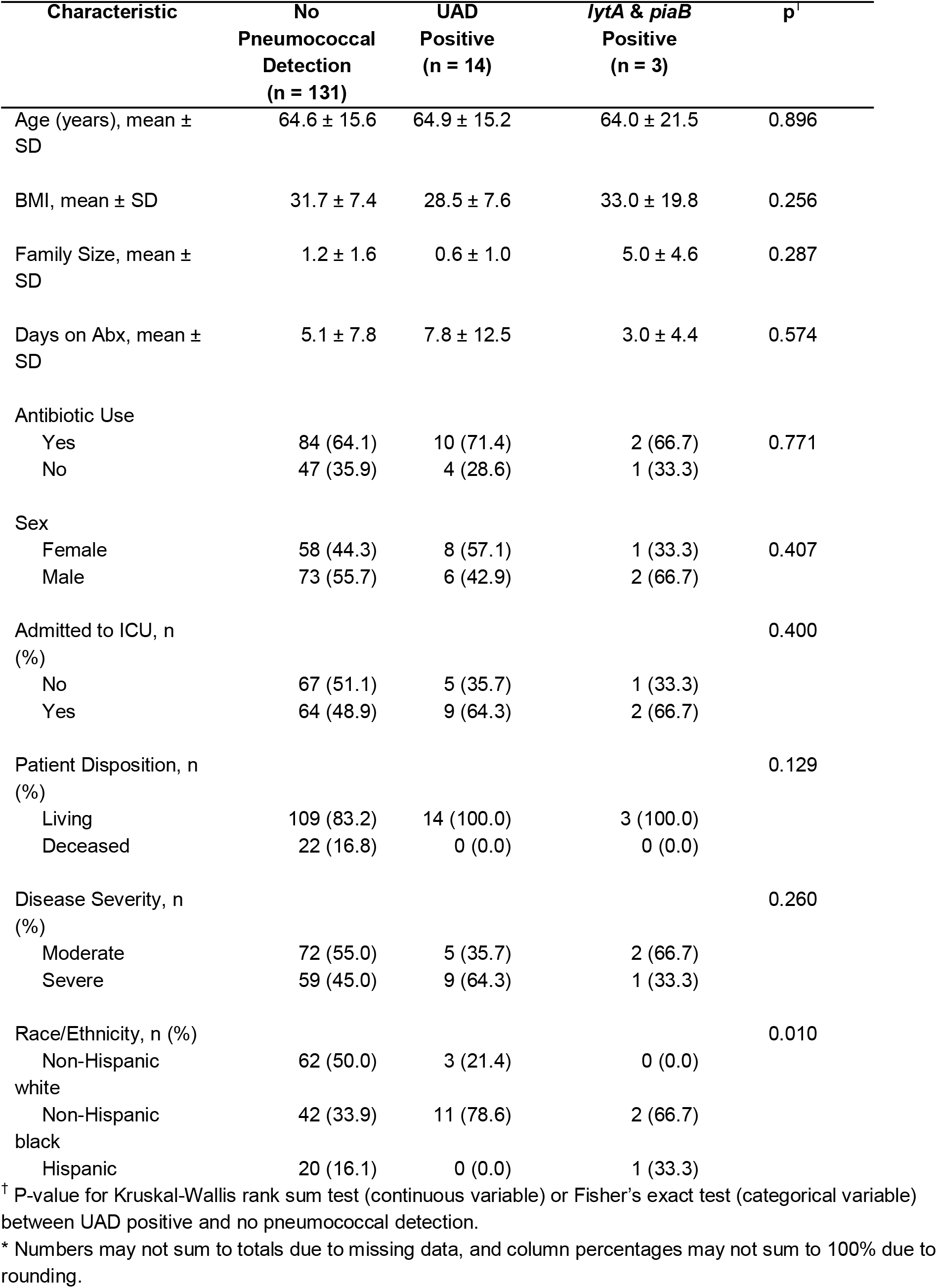
Characteristics of patients categorized by pneumococcal status

### Detection of pneumococcal carriage or pneumococcal LRTI in participants hospitalized with COVID-19

In this study population, 14/127 (11%; SSUAD, n=8; BinaxNOW, n=6) participants tested by UAD had a positive result for at least one sample, indicating pneumococcal LRTI, and 3/96 (3.1%) had a positive saliva-RT-qPCR for at least one sample, indicating pneumococcal carriage (Table 1). None tested positive by both RT-qPCR and urine for pneumococcus. Three individuals returned indeterminate UAD results, which were treated as negative in the remaining analyses (**Figure 1**). For the eight urine samples which tested positive by SSUAD, serotypes detected included 4, 5, 17F, 22F, and 19A in one patient each and 33F in three separate patients. None of the individuals had received a pneumococcal vaccination in the 30 days prior to testing UAD-positive.

**Figure 1.**
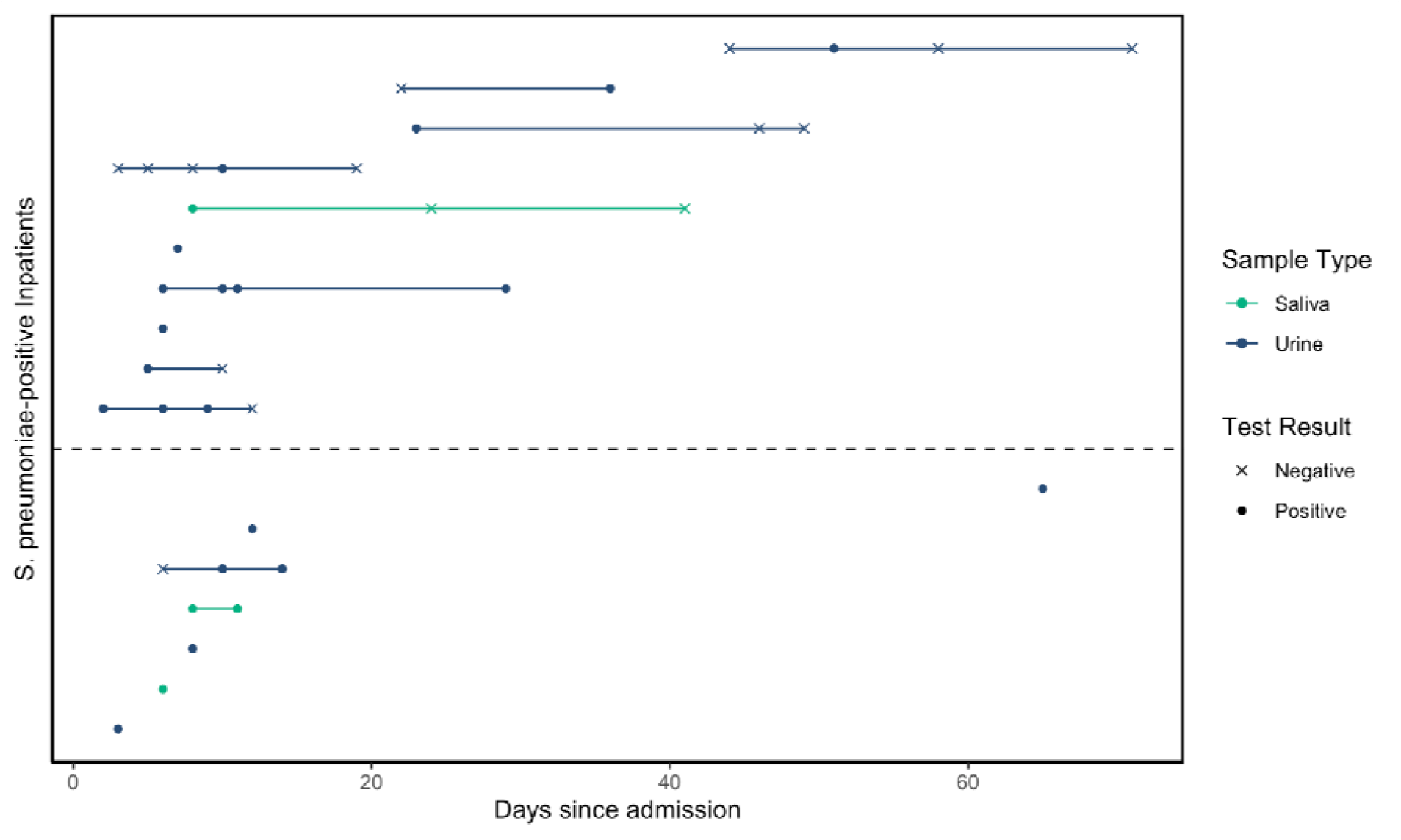
Pneumococcal detection in COVID-19 inpatients. Of the COVID-19 inpatients who tested positive for pneumococcus, detection was exclusively via PCR testing of culture-enriched saliva (indicating carriage; green) or urinary antigen detection (indicating pneumococcal etiology; blue). For some individuals, multiple samples were tested, as shown by points connected by the solid line. Individuals are sorted by time from admission to first positive test for pneumococcus. The dashed line indicates separation of individuals with COVID-19 cases classified as severe (top) and moderate (bottom). All COVID-19 inpatients who tested positive for pneumococcus survived.

### Characteristics associated with detection of pneumococcus

With just 14 positive UAD samples, definitive associations cannot be inferred. However, there were some demographic differences between patients with and without detection of pneumococcus, including larger reported family sizes amongst those with pneumococcal carriage (Table 2). Detection of pneumococcus by both UAD and qPCR was more common in non-Hispanic black patients.

Pneumococcus was detected by UAD more commonly in severe than moderate cases of COVID-19 (Figure 2; OR: 2.20; 95% CI: [0.72, 7.48]). Severe disease was more frequent in non-Hispanic black patients as compared to non-Hispanic white patients (OR: 1.92, 95% CI: [0.93, 4.02]). Death was less common in UAD-positive patients; if the case-fatality rate of 16.8% amongst those patients who did not test positive by UAD were applied to those who were UAD-positive, we would have expected 2-3 (2.7) deaths in this latter group of 14 patients. However, the numbers were small for both severe disease and death with a high degree of uncertainty.

**Figure 2.**
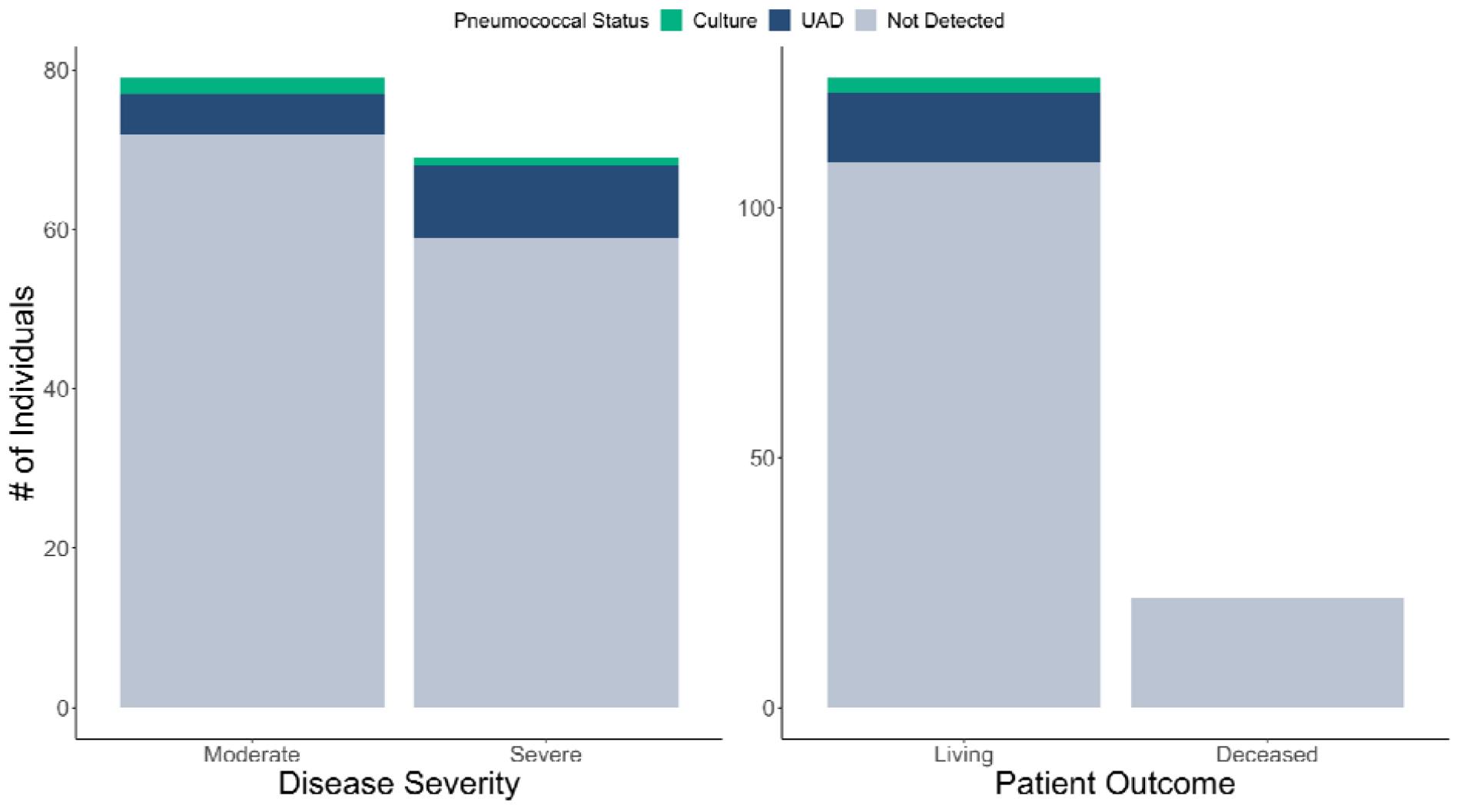
Outcome Measures of COVID-19 infection categorized by pneumococcal status. UAD-positive individuals are disproportionately categorized as severe disease, particularly in comparison to the other pneumococcal statuses. Despite this, deceased individuals had no detection of *S. pneumoniae* and no UAD-positive individuals died while hospitalized for COVID-19.

## DISCUSSION

Based on previously identified interactions between pneumococcus and viral respiratory pathogens, there was concern that pneumococcus and SARS-CoV-2 could synergistically interact and contribute to the burden from the pandemic. With other viral-bacterial co-infections, the primary viral infection induces changes in the host (immune response, integrity of the airway epithelium), and carried bacteria - including pneumococci - which then increases the likelihood that bacteria can both proliferate and cause severe infection.^23^ Moreover, increasing evidence exists suggesting that bacteria can influence host immune responses to viruses.^24,25^ Therefore, we hypothesized that mixed infections between bacteria and viruses could influence the severity of the initial viral infection. To evaluate this hypothesis, we tested saliva and urine samples collected from COVID-19 inpatients in 2020 for markers indicative of the presence of pneumococcal carriage and infection. While detection of pneumococcus in saliva by culture was low relative to previously reported prevalence for non-elderly^16^ and elderly adults in the community,^18^ detection of pneumococcal antigens by UAD (11.0%), was similar to levels previously reported throughout the U.S. amongst older adults with community-acquired pneumonia (CAP).^26^ This, together with similar findings from Italy (13.0%)^27^ and Spain (9%),^28^ suggests that secondary infections in COVID-19 patients may be on par with pre-pandemic rates of detection in CAP.

Despite decreased rates of IPD reported from around the world, LRTI with pneumococcus has still been detected. While the small number of patients made it difficult to draw firm conclusions, pneumococcus was more commonly detected by UAD in individuals with severe COVID-19. This observation may be confounded by the role of race in both increased exposure to pneumococcus and severity of COVID-19 outcomes. Larger datasets that would allow for multivariate analysis would provide additional insights on this question. Further questions can be raised as to whether a UAD-positive CAP (vs. pneumococcal CAP with a false negative UAD) is predictive of disease severity or whether more severe cases of COVID-19 are more likely to develop a secondary pneumococcal pneumonia due to existing disease pathology.

Amongst individuals testing positive for pneumococcus by either culture or UAD, none tested positive by both. PCR testing of culture-enriched saliva is intended for detection of carriage and requires the ability to isolate viable bacteria.^22^ Culture-enrichment of saliva limits detection to living bacteria,^29^ excluding the detection of bacteria killed by antibiotic use. Prior studies suggest that antibiotic use in inpatients may be upwards of 70%.^30^ Our study population had a similar level of antibiotic use (64.9%) at any point during hospitalization for COVID-19. With such a high level of antibiotic usage our estimates of the proportion of pneumonia due to pneumococcus may be underestimated, and may have led to misclassification of pneumococcal status.

Three of eight serotypes detected by SSUAD were serotypes covered by PCV13, with seven of eight contained in PCV15, PCV20, and 23-valent pneumococcal polysaccharide vaccine (PPV23). While PPV23 is not considered to provide substantial protection against non-bacteremic pneumonia by the US CDC,^31^ conjugate vaccines do. If serotype distribution is confirmed by larger studies, new higher-valency PCV15 and PCV20 should provide substantial public health benefit to the older adult population.

One of the major limitations of this study is small sample size. Future studies could be expanded to include more individuals of varying disease severities and demographic and socio-ecologic backgrounds with more complete sample and data collection. This could be done both retrospectively through chart reviews and analysis of previously stored samples or prospectively with new cases and data collection. Increasing longitudinal saliva and urine collection during both inpatient and outpatient stay to monitor both existing and newly acquired carriage as well as secondary infection throughout the duration of disease could help inform important questions on the roles of these different types of pneumococcal states. Furthermore, longitudinal surveillance on both the general community as well as inpatients would best inform viral-bacterial interactions in disease pathogenesis, including the interaction between SARS-CoV-2 and pneumococcus.

In conclusion, these data confirm that patients hospitalized with COVID-19 continued to experience LRTI with *S. pneumoniae* despite dramatic declines in IPD and are suggestive of an association between detection of pneumococcus and severity of COVID-19. Future studies should aim to more deeply and longitudinally explore these dynamics to better assess how pneumococcal carriage may interact with SARS-CoV-2 to influence COVID-19 severity, as well as the degree to which the combination of COVID-19 and pneumococcal infection influence disease outcomes.

## Data Availability

All data produced in the present study are available upon reasonable request to the authors.

## ACKNOWLEDGEMENTS

We thank the study participants for their contribution to our study and the Yale IMPACT biorepository team for their assistance with sample collection.

## AUTHOR’S CONTRIBUTIONS

ALW conceived the study. LRG, RA-P, AA, BDG, DMW and ALW designed the study protocol. AEW, SFF, RA-P, and ALW managed the study. AEW, IMO, DY-C, CAH and ALW were responsible for sample receipt, processing and testing. AEW, LRG, IMO, SBC, DY-C, CAH, MN, SFF, and ALW collected the data. AEW, DMW and ALW performed the analyses and interpreted the data. AEW, DMW, and ALW drafted the manuscript. All authors amended and commented on the final manuscript.

